# Listening to Mom in the Neonatal Intensive Care Unit: A randomized trial of increased maternal speech exposure on white matter connectivity in infants born preterm

**DOI:** 10.1101/2024.09.20.24314094

**Authors:** Katherine E. Travis, Melissa Scala, Virginia A. Marchman, Hua Wu, Cory Dodson, Lisa Bruckert, Molly Lazarus, Rocio Velasco Poblaciones, Kristen Yeom, Heidi M Feldman

**Affiliations:** Division of Developmental-Behavioral Pediatrics, Department of Pediatrics, Stanford University School of Medicine, Stanford, CA, United States; Burke-Cornell Medical Research Institute, Department of Pediatrics, Weill Medical College, Cornell University, New York, NY; Division of Neonatal and Developmental Medicine, Department of Pediatrics, Stanford University School of Medicine, Stanford, CA United States; Department of Psychology, Stanford University, Stanford, CA, United States; Cognitive and Neurobiological Imaging Center, Stanford University, Stanford, CA United States; Department of Radiology, Stanford University School of Medicine, Stanford, CA, United States

## Abstract

**Background:** Early speech experiences have been proposed to contribute to the development of brain structures involved in processing spoken language. However, previous research has been limited to correlational studies. Here, we conducted an RCT with preterm neonates to determine whether increased exposure to maternal speech during NICU hospitalization is causally linked to structural white matter maturation.

**Methods:** We enrolled 46 preterm neonates (24-31 weeks gestational age). Participants were randomly assigned to receive increased (T: n=21) or routine (C: n=25) exposure to mother’s speech. The T-group heard 10-minute audio recordings of their mothers reading a children’s story two times/hour between 10pm-6am, increasing speech exposure by 2.67 hours/day. At near-term-equivalent age, we obtained two high-angular resolution diffusion MRI (scan 1 bvalue=700, scan 2 bvalue=1500) and quantitative T1 relaxometry scans. We assessed mean diffusivity (MD), pre-registered primary outcome (NCT02847689), of the left and right arcuate fasciculus, tracts implicated in language processing.

Secondary outcomes included fractional anisotropy (FA) and R1 (1/T1).

**Findings:** T- and C-groups were equivalent on medical and demographic variables. Compared to the C-group, the T-group demonstrated significantly lower MD in the left (scan 1: mean difference **Δ** *=*0.11, 95% CI:0.03 – 0.19; scan 2: **Δ** *=*0.13, 95% CI:0.04 - 0.21) but not right arcuate (scan 1: **Δ** *=*0.06, 95% CI: -0.23 – 0.15; scan 2: **Δ** *=*0.05, 95% CI:-0.05 - 0.13). The T-group also demonstrated significantly higher FA (scan 1: **Δ** *= -*0.02, 95% CI:-0.04 – -0.00; scan 2: **Δ** *=* -0.03, 95% CI:-0.06 – -0.00) and R1 (**Δ** *=*-0.02, 95% CI:-0.04 – -0.01) in the left but not right arcuate.

**Interpretation:** Preterm neonates who experienced increased exposure to maternal speech during hospitalization demonstrated more mature microstructure of the left arcuate. Findings provide evidence for a causal link between speech experiences and brain development. Increasing speech exposure in the NICU may benefit preterm children.

**Research in Context Panel:** *Evidence before this study:* Observational studies document the importance of early speech experience for language learning and brain development in term and preterm children. Children born preterm are at-risk for adverse language outcomes that have been attributed to alterations in brain development from limited exposure to speech in the neonatal intensive care unit (NICU). However, evidence that early speech experiences causally effect the development of brain structures relevant for language is lacking.

*Added value of this study:* The Listening to Mom in NICU study is the first randomized controlled trial specifically designed to test the causal effects of maternal speech exposure on white matter brain development in neonates born preterm. This study demonstrates that speech experiences during neonatal development directly contribute to the maturation of the left arcuate fasciculus, a white matter tract implicated in language.

*Implications of all the available evidence:* Study findings advance understandings for how early speech experiences contribute to neonatal brain development. This study also demonstrates that increasing exposure to speech via audio recordings among infants born preterm could serve as an inexpensive and scalable intervention to support recovery from alterations in brain development related to the NICU experience.

## Introduction

Early speech experiences are critical for language learning and are thought to contribute to the development of brain structures involved in processing spoken language (1). However, evidence of causal linkages are currently lacking. Children born very preterm (< 32 weeks gestational age) are at risk for experiencing delays in language development and subsequent language-based learning disabilities (2). Adverse language outcomes in children born preterm have been linked to alterations in structural brain development (3,4), particularly changes in the microstructure of white matter circuits (5). Brain development in preterm children may be affected by experiential factors, such as reduced exposure to maternal speech from spending multiple weeks to months in the neonatal intensive care unit (NICU)(6,7). In-utero, fetuses experience 2-3 times more speech from their mothers than preterm infants at equivalent developmental ages (8). Interventions designed to enrich speech experiences in the NICU have shown positive benefits to short-term health outcomes among neonates born preterm (9). Studies have yet to establish whether modifying speech experiences in the NICU can impact brain development in structures directly linked to language abilities in children born preterm.

Here, we peformed a randomized controlled study to assess causal links between an intervention to increase exposure to spoken language and development of white matter circuits in infants born very preterm. To supplement speech experiences in the NICU, babies in the treatment group were played audio recordings of their mothers reading a children’s storybook and babies in the control group did not hear the audio recordings. We assessed intervention effects on measures of white matter microstructure acquired with diffusion MRI scans at near-term-equivalent age, prior to hospital discharge. We chose mean diffusivity (MD), a metric from dMRI, as the primary outcome measure given previous evidence that found MD to be sensitive to age-related changes during development (10,11). We focused on white matter tracts of the left and right arcuate fasciculus. These white matter pathways, and particularly the left arcuate, are known to be involved in speech and language processes (12). We hypothesized that neonates randomized to the treatment group would show decreased MD within the arcuate, evidence of increased maturation, compared to neonates in the control group. Such findings would advance our understandings of how early experiences contribute to brain development. In addition, if successful, increasing exposure to speech via audio recordings among infants born preterm could serve as an inexpensive and scalable intervention to support recovery from alterations in brain development related to the NICU experience.

## Methods

### Study design

This study was a prospective, parallel-group, randomized control trial, performed in a single neonatal intensive care nursery at the Lucile Packard’s Children’s Hospital (LPCH) Stanford University in Palo Alto, California. The study protocol was was approved by the Stanford School of Medicine Institutional Review Board and registered at Clincaltrials.gov with protocol description (NCT02847689).

### Participants

Female and male infants were eligible to enroll if they were born between 24-0/7 and 31-6/7 weeks gestational age (GA). We recorded sex, gestational age at birth, and birthweight as indicated in the electronic medical record. Infants were excluded from participation if any of the following criteria were met: (1) congenital anomalies or recognizable malformation syndromes, (2) serious neurological conditions, including active seizure disorders, history of central nervous system infections or hydrocephalus, intraventricular hemorrhage grades III-IV, or cystic periventricular leukomalacia, (3) surgical treatment of necrotizing enterocolitis, (4) small for gestational age (< 3^rd^ percentile) and/or intra-uterine growth restriction, (5) twin-to-twin transfusion, (6) likelihood to be transferred from LPCH NICU to alternate care facility prior to 36 weeks postmenstrual age or the pre-discharge MRI scan. (7) major sensori-neural hearing loss.

Families of eligible infants were approached once an infant was determined by clinical staff to be medically stable, which typically coincided with infants’ transition to a step-down unit, the Packard Intermediate Care Nursery (PICN). Parents of enrolled participants gave written informed consent before randomization.

### Randomization

Enrollment was performed by a study research coordinator, with assistance from the principal investigator (KET). Participants were randomly allocated to either the Treatment (T, speech exposure) or the Control (C, standard of care) group by the principal investigator using a minimization algorithm by Pocock and Simon (13) implemented in the R statistical software package. Randomization was stratified for gestational age at birth (24-0/7 to 27-6/7 weeks or 28-0/7 to 31-6/7 weeks), to control for potential developmental differences in response to the intervention, and for socioeconomic status (SES) (low versus high SES; indexed as income status from public versus private insurance) to control for potential differences in language outcomes affected by SES factors. Twins and multiples were assigned to the same group. Families and clinical staff were not informed of group status.

### Procedures

#### Speech Recordings

Following consent and prior to randomizaion, all mothers were audio-recorded while reading aloud the first chapter from the children’s storybook, *Paddington Bear* (14), in their native language. Recordings were subsequently post-processed to have the same sound intensity (45dB) and divided into two 10- minute segments using the auditory software Praat (http://www.fon.hum.uva.nl/praat/).

#### Delivery of Intervention

We played audio recordings of maternal speech for infants randomized to the T-group at hourly intervals overnight (10pm – 6am). This time window coincided with periods of relative quiet in the intermediate care unit, when families were unlikely to visit and provide natural speech input to the child. Within a given hour, 10-min segments were played at prespecified intervals separated by at least 20 minutes to avoid synchronization with biological and sleep rhythms. Infants in the T-group thus heard a total of approximately 2.67 h (160 minutes) of speech recordings per night (20 min/h × 8 h), far above the average of 20-50 minutes, the usual speech exposure of infants cared for in open-bay NICU settings (15). Recordings were played automatically via a timer function on an iPod to minimize involvement of clinical staff and to ensure regular timing of recordings. To keep families and staff from determining group status, all participants had an iPOD placed in their crib or isolette near their head regardless of group assignment. Sound intensity for speech recordings was played below hourly safety levels < 50 dB (5). Infants randomized to the C-group had their mothers record themselves reading and had the same the auditory setup placed in their cribs (ie., iPod); however, no recordings were played. Duration of intervention was defined as number of nights between when first nightly recording began or mock auditory set-up was first placed in a crib to the date of MRI scan, which typically preceded hospital discharge (ie., approximately 36–38 weeks post-menstrual age (PMA)). The percentage of days parents visited during the intervention period was also measured to approximate balance in caregiver speech between groups.

#### Clinical MRI scanning and sequence parameters

At the time of the study, standard of care at LPCH for all infants born very preterm (<32 weeks gestational age) included MRI scans at near-term-equivalent age, prior to hospital discharge. Per clinical procedures, infants were scanned during natural sleep without the use of sedation. For this study, infants were scanned on a 3-T MRI (GE-Discovery MR750) equipped with either an 8-channel HD head coil (n=28) or a 32-channel (n=5) HD head coil (General Electric Healthcare, Little Chalfont, UK). Scans were collected by hospital technologists unaware of group assignment. For purposes of this study, we collected a high-angular resolution (60-direction) diffusion MRI sequence scan 1: b=700 s/mm^2^ with multi-slice echoplanar imaging (MS-EPI) for rapid image acquisition and two-six volumes b=0. An additional high-angular resolution (60-direction) diffusion MRI sequence scan 2: b=1500 s/mm^2^ was included in the clinical neuroimaging protocol and used to validate findings from scan 1. A quantitative T1 relaxometry scan, also part of the clinical protocol, was performed with a slice-shuffled, inversion recovery EPI (IR-EPI) sequence with multiple inversion times (TI) and a second IR-EPI with the reverse-phase encoding direction. qT1 relaxometry scans were analyzed to validate and to interpret dMRI findings. Both dMRI and qT1 sequences were collected with a spatial resolution of 2.0mm^3^ with full brain coverage. The clinical protocol included a high-resolution T1-weighted scan (1mm^3^ voxel size) that was used as the participant-specific anatomical reference for analyses of dMRI data.

#### Neuroimaging pre-processing and tractography analysis

MRI data were managed and analyzed using a cloud-based neuroinformatics platform (Flywheel.io). Procedures our team developed for diffusion MRI pre-processesing and tractrography analysis of neonatal clinical MRI scans are described in Dubner et al (2023)(16). Details for the these procedures are provided in the supplemental information along with additional procedures for qT1 preprocessing and analyses (see Supplemental information). These procedures were used to obtain the primary outcome measure, mean diffusivity (MD) and secondary outcome measures, fractional anisotropy (FA) and relaxtion rate (R1 sec^-1^) from the left and right arcuate fasciculus tracts. Figure 2 shows tractograms of the left and right arcuate fasciculus generated from each of the two dMRI scans (bvalue=700 and bvalue=1500) and displayed on the T1w image from a single participant.

### Outcomes

The primary outcome was white matter mean diffusivity (MD) of the left and right arcuate fasciculus. Post-hoc secondary outcome measures included fractional anisotropy (FA) from dMRI and relaxation rate R1 (R1=1/T1) from quantitative T1 relaxometry scans also measured from the left and right arcuate fasciculus.

The present intervention was considered minimal risk. Study monitoring was assessed by the PI, the institutional review board and an internal data safety monitoring board comprised of study consultants, including board certified neonatologists and a pediatric neuroradiologist at Stanford Medical School. No adverse events were reported.

### Statistical Analysis

We planned enrollment of n=42 participants (21 per group) based on power calculations that used data from an existing diffusion MRI intervention study of preterm infants (17). Planned enrollment was expected to have power of ß = 0.88 to detect significant group differences of large effect size (Cohen’s *d* = 1.0). All statistical analyses were performed in R (version 4.3.1). We used an intention to treat strategy for all analyses. Welch t-tests were used to assess group differences between treatment and control groups on the primary outcome measure given differences in sample size. Secondary analyses performed to confirm treatment effects also used Welch t t-tests to assess group differences in fractional anisotropy (FA) from dMRI scans and relaxation rate (R1) from the quantitative T1 relaxometry scan. We report the magnitude of group differences as the absolute mean difference **Δ** with corresponding 95% confidence intervals and the effect size of group differences as Cohen’s d. Exploratory analyses used Pearson correlations to examine associations between the primary outcome metric (MD) and additional white matter metrics (FA, R1) used to confirm treatment effects. Statistical significance was set at p<0.05. No external data monitoring committee was used for this study. Trial is listed as Listening to Mom in the NICU: Neural, Clinical and Language Outcomes # NCT02847689 on clinicaltrials.gov.

### Role of the funding source

The funder of the study had no role in study design, data collection, data analysis, data interpretation or writing of the report.

## Results

Between September of 2016 and May of 2019, a total of 46 healthy neonates born very preterm were consented and randomized to either the treatment group (n=21) or control (n=25) groups (Figure 1). From the initial sample, 33 neonates (n =19 T group; 14 C group) had diffusion MRI scans available for analysis. Baseline clinical, demographic and intervention characteristics were matched between groups in both the initial randomized sample (Table 1) and in the final sample with available dMRI data (n=33) (Table S1).

**Figure 1.**
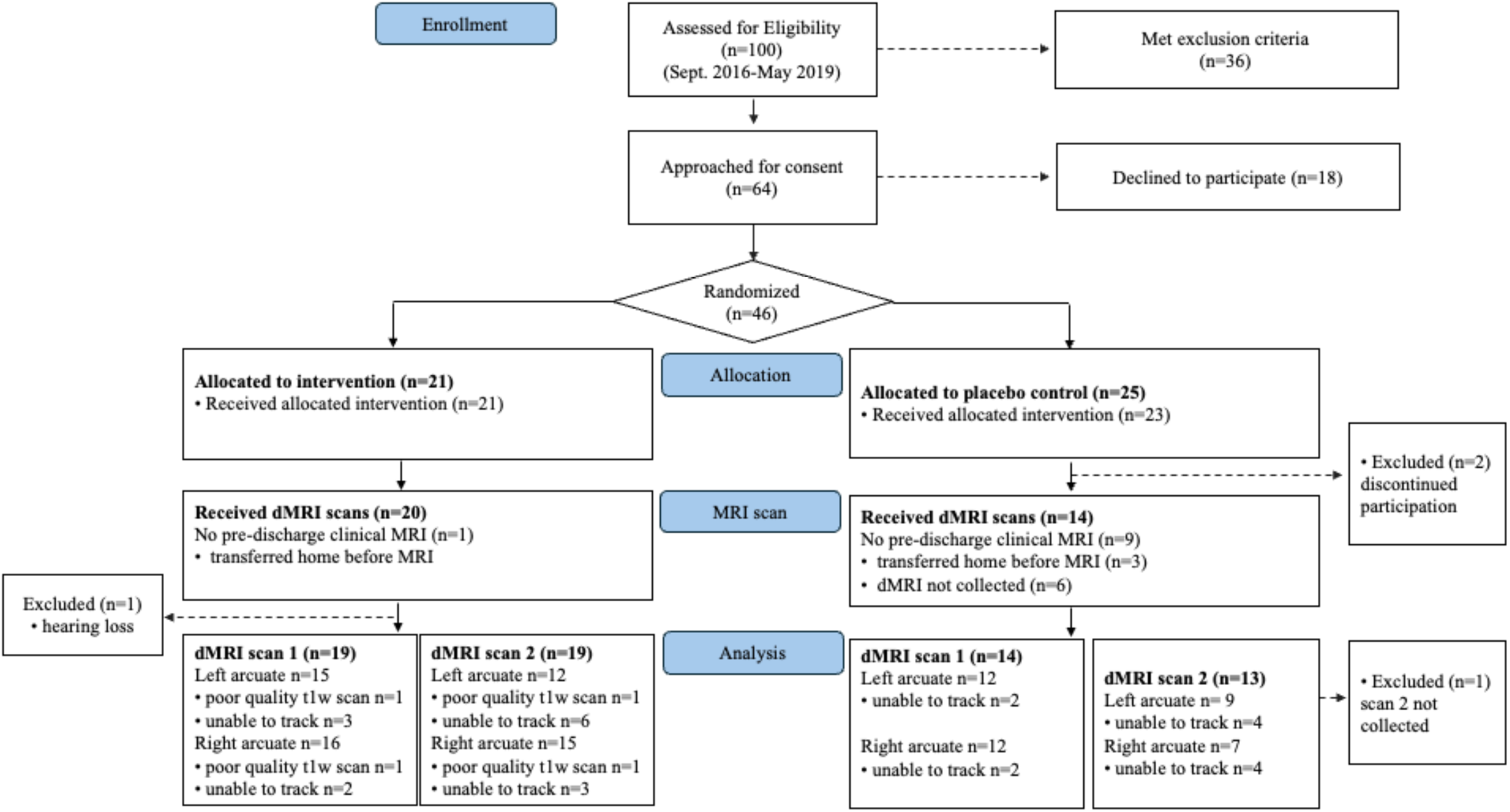
Consort Diagram

**Figure 2.**
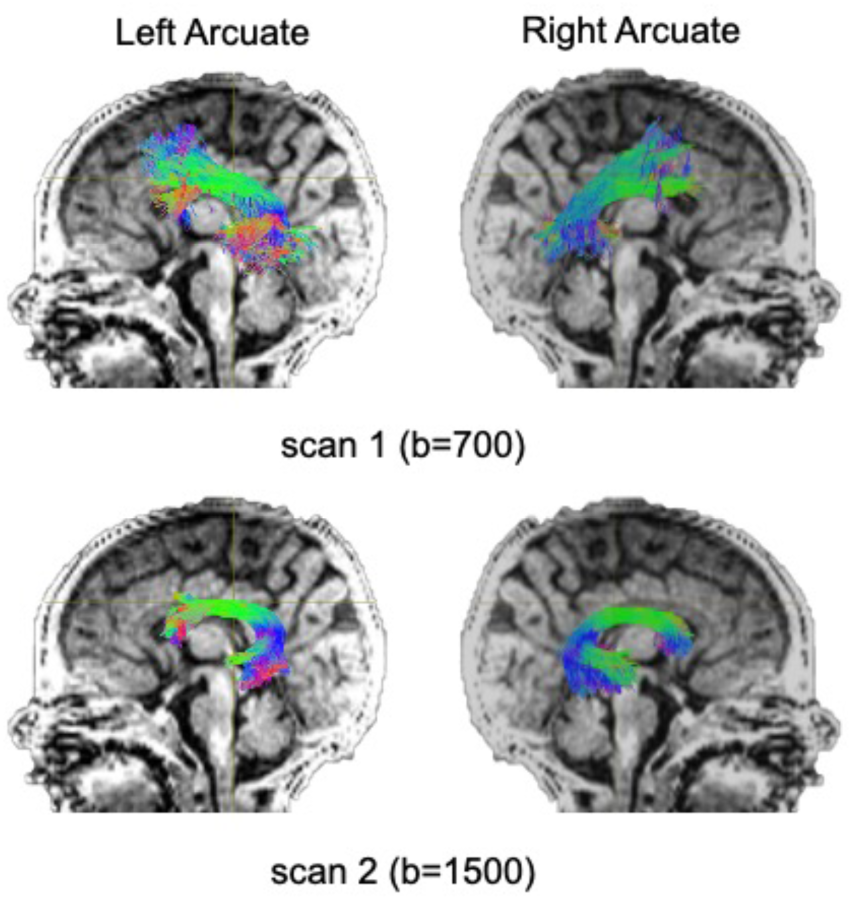
Tract renderings illustrate the left and right arcuate fasciculus from a single infant participant born very preterm. Tract renderings from each of two separate diffusion MRI scans (scan 1, bvalue = 700 or bvalue 1500) are displayed on a mid-saggital T1-weighted (T1w) image. Colors represent primary orientation of streamlines (red = left-right, green = anterior-posterior, blue = superior-inferior).

**Table 1.**
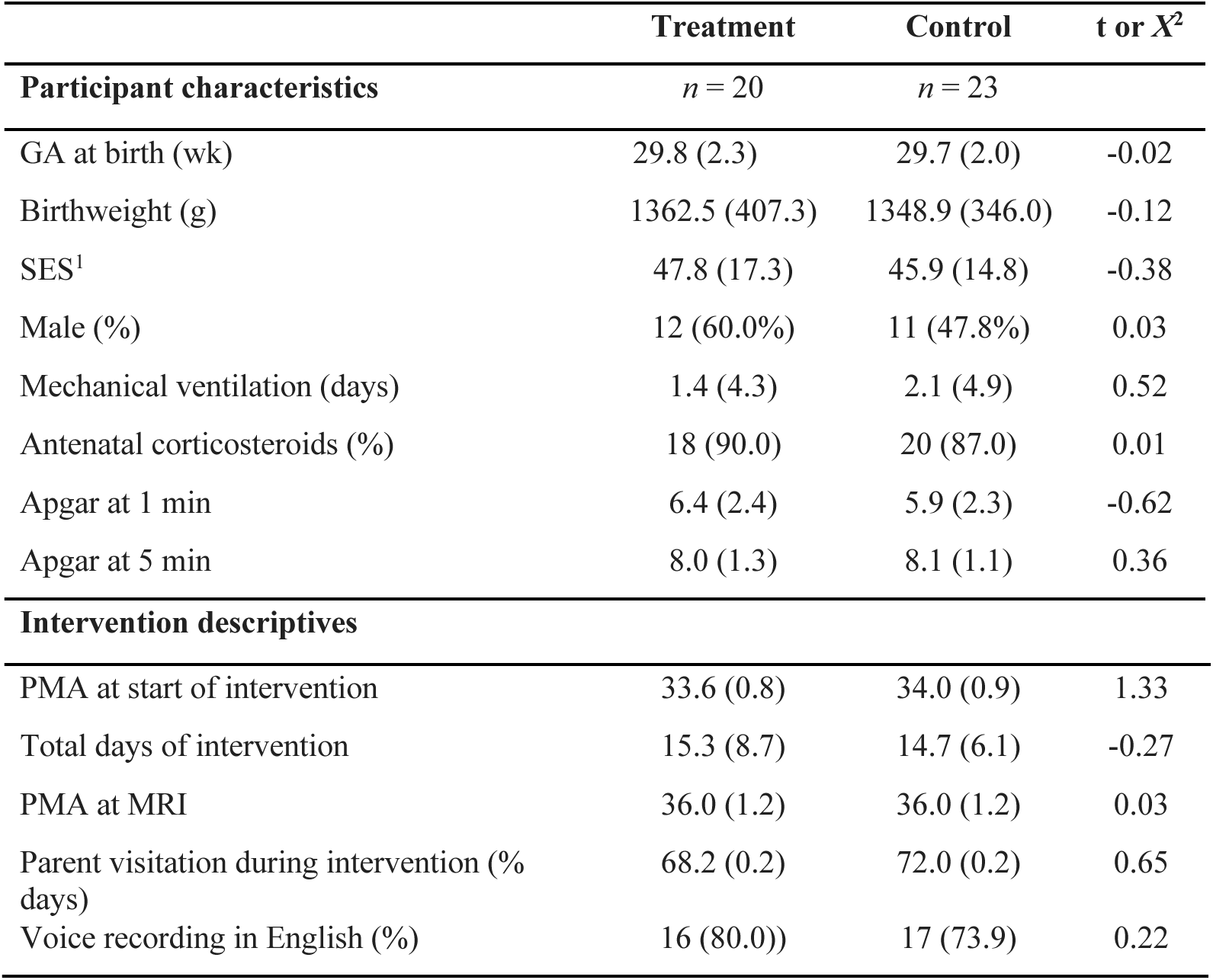
Trial intervention participant characteristics and intervention descriptives.

| | Treatment | Control | t or $X^2$ |
| --- | --- | --- | --- |
| Participant characteristics | *n* = 20 | *n* = 23 |  |
| GA at birth (wk) | 29.8 (2.3) | 29.7 (2.0) | -0.02 |
| Birthweight (g) | 1362.5 (407.3) | 1348.9 (346.0) | -0.12 |
| SES<sup>1</sup> | 47.8 (17.3) | 45.9 (14.8) | -0.38 |
| Male (%) | 12 (60.0%) | 11 (47.8%) | 0.03 |
| Mechanical ventilation (days) | 1.4 (4.3) | 2.1 (4.9) | 0.52 |
| Antenatal corticosteroids (%) | 18 (90.0) | 20 (87.0) | 0.01 |
| Apgar at 1 min | 6.4 (2.4) | 5.9 (2.3) | -0.62 |
| Apgar at 5 min | 8.0 (1.3) | 8.1 (1.1) | 0.36 |
| Intervention descriptives |  |  |  |
| PMA at start of intervention | 33.6 (0.8) | 34.0 (0.9) | 1.33 |
| Total days of intervention | 15.3 (8.7) | 14.7 (6.1) | -0.27 |
| PMA at MRI | 36.0 (1.2) | 36.0 (1.2) | 0.03 |
| Parent visitation during intervention (% days) | 68.2 (0.2) | 72.0 (0.2) | 0.65 |
| Voice recording in English (%) | 16 (80.0)) | 17 (73.9) | 0.22 |

Diffusion MRI tractography analyses was successful in identifying the left and right arcuate in the majority of infants for both scans (scan 1: left 82%; right 85%; scan 2: left 66%; right 69%). Table 2 presents group means for primary (MD) and secondary (FA, R1) outcome measures and results of statistical analyses performed to assess group differences. Figure 3 presents box plots to visualize group comparisons for MD and FA measured from both dMRI scans (scan 1 and scan 2) and R1 measured from the qT1 scan.

**Figure 3.**
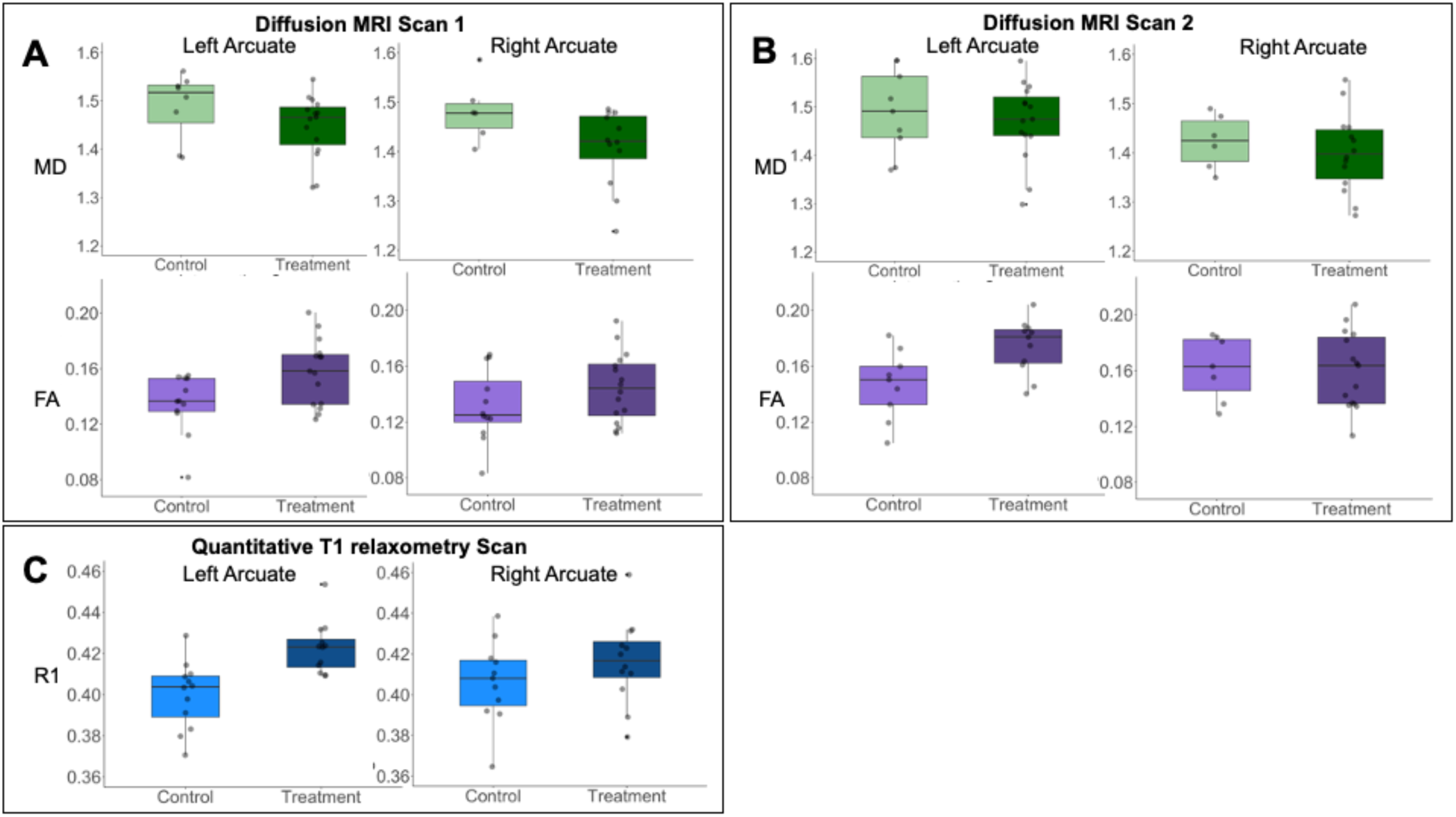
Box plots demonstrate group comparisons for primary and secondary outcome measures used to assess white matter microstructure. Panel A demonstrates comparisons between treatment and control groups for mean diffusivity (MD, primary outcome) and fractional anisotropy (FA, secondary outcome) obtained from the left and right arcuate from diffusion scan 1 (bvalue = 700). Panel B demonstrates comparisons between treatment and control groups for mean diffusivity (MD, primary outcome) and fractional anisotropy (FA, secondary outcome) obtained from the left and right arcuate from diffusion scan 2 (bvalue = 1500). Panel C demonstrates group differences between treatment and control groups for secondary outcome measure relaxation rate (R1) obtained from the left and right arcuate from the quantitative T1 relaxometry scan. Box plots illustrate median with first and third quartiles. Vertical lines illustrate 2.5-95%iles. Gray scatter dots represent individual participants. Treatment group is indicated by dark colors and the control group is indicate by light colors for each white matter outcome metric (MD, FA, R1)

**Table 2.** Group comparisons for primary and secondary outcomes of white matter microstructure.

|  | Treatment | Control |  |  |  |
| --- | --- | --- | --- | --- | --- |
| Primary outcome: MD | Mean (std), n | Mean (std), n | *t* | Mean Difference $\Delta$<br>(95% Confidence Interval) | Effect Size<br>(*d*) |
| **Scan 1** |  |  |  |  |  |
| Left Arcuate Fasciculus | 1.44 (0.06), 15 | 1.56 (0.11), 12 | 2.90 (p=0.01) | 0.11 (0.03 – 0.19) | 1.35 |
| Right Arcuate Fasciculus | 1.48 (0.09), 16 | 1.54 (0.12), 12 | 1.53 (p=0.14) | 0.06 (-0.23 – 0.15) | 0.57 |
| **Scan 2** |  |  |  |  |  |
| Left Arcuate Fasciculus | 1.41 (0.08), 12 | 1.54 (0.10), 9 | 3.26 (p=0.005) | 0.13 (0.04 - 0.21) | 1.44 |
| Right Arcuate Fasciculus | 1.42 (0.10), 15 | 1.45 (0.09), 7 | 0.84 (p=0.41) | 0.05 (-0.05 - 0.13) | 0.31 |
| **Secondary outcome: FA** |  |  |  |  |  |
| **Scan 1** |  |  |  |  |  |
| Left Arcuate Fasciculus | 0.16 (0.02), 15 | 0.13 (0.02), 12 | -2.62 (p=0.01) | -0.02 (-0.04 – -0.00) | 1.50 |
| Right Arcuate Fasciculus | 0.14 (0.02), 16 | 0.13 (0.03), 12 | -1.34 (p=0.19) | -0.02 (-0.03 – 0.01) | 0.39 |
| **Scan 2** |  |  |  |  |  |
| Left Arcuate Fasciculus | 0.18 (0.02), 12 | 0.15 (0.02), 9 | -2.96 (p=0.01) | -0.03 (-0.06 – -0.00) | 1.50 |
| Right Arcuate Fasciculus | 0.16 (0.03), 15 | 0.16 (0.02), 7 | 0.15 (p=0.88) | 0.00 (-0.02 – 0.03) | 0.00 |
| **Secondary outcome: R1** |  |  |  |  |  |
| **Scan 3** |  |  |  |  |  |
| Left Arcuate Fasciculus | 0.42 (0.01), 12 | 0.40 (0.02), 12 | -3.80 (p=0.001) | -0.02 (-0.04 – -0.01) | 1.26 |
| Right Arcuate Fasciculus | 0.42 (0.02), 12 | 0.41 (0.02), 11 | -1.18 (p=0.25) | 0.01 (-0.03 – 0.01) | 0.50 |

MD = mean diffusivity ( $\mu^2/\text{ms}$ ), FA= fractional anisotropy, R1 = relaxation rate (1/seconds)

Analysis of MD, the primary outcome measure, obtained from the arcuate tracts revealed that neonates in the T-group demonstrated significantly lower MD compared to neonates in the C-group in the left but not the right arcuate in both dMRI scans (Table 2, Figure 3). Secondary post-hoc analyses of FA from dMRI scans and R1 from the quantitative T1 relaxometry scan confirmed treatment effects within the left arcuate fasciculus, with neonates in the treatment group demonstrating significantly higher FA and higher R1 compared to neonates in the control group (Table 2, Figure 3). No effect of treatment was observed within the right arcuate for either FA or R1 (Table 2, Figure 3). Exploratory analyses revealed that all metrics of white matter microstructure were associated in expected directions. Specifically, MD was observed to be strongly and negatively correlated with both R1 and FA in the left and right arcuate, while FA and R1 showed moderate positive correlations in the left and right arcuate (Supplementary Figure S1).

## Discussion

This randomized control trial established the effect of increased speech exposure during NICU hospitalization on brain development in healthy infants born very preterm. Compared to infants in the control group, infants who experienced increased exposure to maternal speech demonstrated evidence for more mature microstructure of the left arcuate fasciculus, a white matter pathway known to be important for language. Importantly, group differences were robust, replicating across three separate MRI scans (two diffusion sequences and quantitative T1 relaxometry) and three measures of white matter microstructure (MD, FA and R1). These findings are novel because they go beyond previous correlational studies to causally link speech experiences during neonatal development to the maturation of brain structures implicated in language. Moreover, these findings suggest that enriching the speech environment of the NICU may facilitate the healthy maturation of white matter pathways, offering important benefits to brain development in children born preterm.

Consistent with our hypotheses, neonates in the treatment group demonstrated significantly lower MD compared to neonates in the control group. This finding aligns with previous studies showing reductions in MD and/or increases in FA in response to language-based interventions in children (18,19), or in response to parent-based NICU interventions for preterm newborns (17). Exploratory post-hoc analyses of our data found strong negative correlations between our primary outcome measure, MD, and R1, a metric from qT1 relaxometry, that has been shown to be highly predictive of myelin content in histological studies (20). Increases in myelin content and reductions in water content from tissue growth are characteristic of changes occurring prenatally during the third trimester and post-natally through the 2^nd^ year of life (21,22). These developmental changes in tissue properties are detectable with the white matter microstructural metrics assessed here (MD, FA, and R1) (20,23) and are presumed to reflect myelination (11). Thus, our findings provide evidence that the intervention led to differences in tissue properties that reflect enhanced white matter maturation.

The large effect size is notable. Effect sizes of similar magnitudes have been reported for dMRI metrics, such as FA, comparing term to very preterm infants at term-equivalent ages (24). The amount of additional speech provided in the treatment condition was comparable to the difference between maternal speech exposure in utero during the third trimester and that experienced by preterm neonates of equivalent ages ex-utero (8). Our ability to detect large effects may thus be because the current intervention closely approximated daily levels of speech exposure necessary for brain development.

Further randomized trials are needed to determine the specific dosing and timing of speech exposure needed to remediate gaps in speech exposure that can occur during NICU hospitalization and that may adversely impact brain development (25). Additional studies are also required to understand if there may be differential effects of live versus recorded caregiver speech on neonatal brain development.

Treatment effects were strongest in the left arcuate and were not significant in the right arcuate. Several lines of evidence suggest that brain areas in both the left and right hemispheres, including the arcuate, are important in processing speech information (12,26,27) and in the development of language (28). Our data provide intriguing evidence that left-hemispheric predominance for language reflects experience-dependent processes that may begin prior to birth. Larger studies are needed to establish whether these intervention effects may be specific to particular brain areas or may influence white matter development more generally. We did not test other pathways given the sample size.

Most evidence to date linking early speech experiences to structural brain development derives from observational studies in older infants (29). To our knowledge, the current findings are the first to isolate speech experiences as having a direct effect on neonatal brain development. Prior randomized controlled trial studies of premature newborns have demonstrated effects of other auditory interventions on structural or functional brain development (30,31). Together with these studies, the current findings underscore the sensitivity of the neonatal brain to auditory-speech experiences. Understanding whether different types of auditory experiences have similar, differential, or combinatorial benefits to neonatal brain development requires further investigation. The present data provide evidence that structural white matter measures used here may have utility as biomarkers for assessing the effects of other neonatal interventions on brain development (6,7).

Auditory therapies and speech interventions may be effective in promoting brain development in preterm newborns possibly by reducing stress or by providing experience-dependent activity to promote neuronal functioning (25,32). Recorded maternal speech in the NICU may also benefit sleep in preterm newborns, a factor relevant for brain development and which may have also been affected by the current intervention (33). Relating measures of structural brain development to physiological or clinical assessments of neurological function will be important for interrogating the neural mechanisms of early NICU auditory interventions. Identifying the underlying neurobiological correlates of such interventions will also benefit from translational research studies in non-human animals.

The current study reveals that clinical interventions may serve as an important experimental model in which developmental studies can interrogate how environmental experiences directly contribute to brain development, possibly for the benefit of behavioral, cognitive or physical outcomes in children.

Interventions designed to enhance caregiver speech exposure may provide opportunities to educate parents about the importance of talking and reading to their babies beyond the NICU. Playing recordings of caregiver speech to infants may also provide opportunites to promote equity in the amounts of caregiver speech experienced by preterm newborns in the NICU, especially for infants from socio-economically disadvantaged backgrounds whose families may not have the financial resources or social supports (e.g., paid work leave, transportation costs, childcare assistance) to be able to visit their infant in the NICU regularly.

This study has limitations. The sample size was small, although adequately powered to detect large effects. Our replication of treatment effects across three separate scans and across three measures provides confidence in our conclusions. We did not have baseline scans to establish whether differences in neural structures predated intervention. This concern is minimized because randomization resulted in groups with comparable medical and demographic profiles in factors that could influence neonatal brain development, such as gestational age and postmenstrual age at scan. While we did not have a direct measure of caregiver speech at bedside, parental visitation rates were comparable between groups and similar to published rates at our center (34). This cohort was not representative of the full range of infants born very preterm, as our sample was limited to relatively healthy preterms. Larger studies would be required to establish efficacy of our intervention for preterm children with more complex medical histories who may be at most risk for alterations in brain development from long hospital stays, inflammatory conditions, and other complications of preterm birth. At the same time, this design feature is also a strength because our findings may more easily generalize to models of brain development in typical developmental circumstances.

In sum, increasing exposure to mother’s speech via overnight audio recordings was directly linked to maturation of the left arcuate fasciculus, a white matter circuit implicated in life-long language processing. The study provided a scientifically necessary causal link between language experience and brain structure. These findings also offer key insights regarding how interventions involving increasing exposure to speech may offer important neuroprotective experiences that can mitigate the effects of prematurity on brain development.

## Data Sharing Statement

De-identified individual participant data that correspond to the results reported in this article will be provided immediately following publication, including analytic code for statistical analyses and figure generation.

## Notes

**Conflict of Interest Disclosure:** The authors declare no conflict of interest.

**Funding/Support:** This research work was supported by grants from the Eunice Kennedy Shriver National Institute of Child Health and Human Development (K.E. Travis, PI: 5R00-HD84749; H.M. Feldman, PI: 2R01-HD069150).

### Competing Interest Statement

The authors have declared no competing interest.

### Clinical Trial

NCT02847689

### Clinical Protocols

https://clinicaltrials.gov/study/NCT02847689

### Funding Statement

This research work was supported by grants from the Eunice Kennedy Shriver National Institute of Child Health and Human Development (K.E. Travis, PI: 5R00-HD84749; H.M. Feldman, PI: 2R01-HD069150).

### Author Declarations

The study protocol was was approved by the Stanford School of Medicine Institutional Review Board and registered at Clincaltrials.gov with protocol description (NCT02847689).

