## Supplemental Materials for "Listening to Mom in the Neonatal Intensive Care Unit: A randomized trial of increased maternal speech exposure on white matter connectivity in infants born preterm"

**Supplemental Information**

**Image Preprocessing**

**Diffusion MRI**

Analyses of neuroimaging dMRI data was implemented in Reproducible Tract Profiles (RTP) (https://github.com/vistalab/RTP-pipeline) (1,2). Diffusion preprocessing, modeling and tractography within RTP is largely based on open-source software from FSL (https://fsl.fmrib.ox.ac.uk/fsl/fslwiki), MRTrix3 (https://www.mrtrix.org/) and Automated Fiber Quantification (AFQ, https://github.com/yeatmanlab/AFQ) (3). A full list of software dependencies can be found here (https://github.com/vistalab/RTP-pipeline/wiki). Procedures for preprocessing were the same for the two diffusion scans collected at different b-values (b=700 or b=1500).

RTP consists of three main steps: (i) Structural processing and Region of Interest (ROI) creation (ii) dMRI preprocessing (RTP-preproc) and (iii) whole-brain tractography and tract segmentation (RTP-Pipeline). Details of these steps are described by Lerma-Usabiaga et al.(2) and Liu et al. (1). We modified the first step of the RTP pipeline to account for the immature neonatal brain. These adaptations included using infant Freesurfer (https://surfer.nmr.mgh.harvard.edu/fswiki/infantFS) (4) for T1-weighted image segmentation and replacing the adult MNI template and ROIs with a neonatal template from the Edinburgh Neonatal Atlas (ENA33) (5) and corresponding neonatal ROIs to enhance ROI placement and tract identification. The brainmask generated by infant freesurfer for each participant was inspected, and in some cases, manually edited using mrTrix to ensure full-brain coverage.

We performed the following diffusion image preprocessing steps (RTP-preproc): (i) data denoising using principal component analysis (6,7), ii) Gibbs ringing correction (8), (iii) eddy current and motion correction (9), and (iv) anatomical alignment of diffusion data to the average non-diffusion-weighted volumes, which were registered to the infant's high-resolution, ac-pc aligned T1-weighted anatomical image using rigid body transformation.

**Quantitative T1 relaxometry**

We used IR-EPI data to estimate relaxation rate R1 (R1 = 1/T1) in each voxel. This was achieved using t1fit (https://github.com/cni/t1fit) software implement in Flywheel. This software includes routines to (1) unshuffle slices within a volume of differing inversion times from slice-shuffled pulse sequence (2) perform epi-distortion correction using FSL topup tool (10,11) and (3) perform T1 fitting using algorithms based on those described in Barral et al. (2010) (12). Output of the t1fit software is the estimated T1 in each voxel. qT1 maps were then aligned to dMRI b=0 maps from scan 1 (bvalue=700), using rigid-body alignment performed using ANTs (13) implement in Flywheel. qT1 maps were visually inspected for alignment and included as inputs to RTP-Pipeline to obtain T1 values for the left and right arcuate. We calculated (R1 = 1/T1) for each tract using mean T1 generate for each tract. We chose to report R1 for ease of interpretation given that higher values of R1 are associated with higher levels of tissue properties related to myelin (14).

**Tractography Analyses**

The preprocessed dMRI data output from RTP-preproc served as the input for diffusion metrics modeling, whole-brain tractography, and tract segmentation in RTP-pipeline. Aligned qT1 maps were supplied as inputs implemented in RTP2 (15). White matter diffusion metrics (FA and MD) were calculated based on the diffusion tensor model. Constrained spherical deconvolution model (CSD) (16) with eight spherical harmonics (lmax = 8) was used to calculate fiber orientation distributions (FOD) for each voxel. To account for the immaturity of the neonatal brain, the FA mask threshold was set to 0.15, and the FOD threshold to 0.08 for CSD-based tractography. The CSD- based tractography consisted of (i) Ensemble Tractography (17) to estimate the whole-brain white matter connectome. MRtrix3 generated three candidate connectomes with varying angle parameters and lengths (1: angle 45°, lengths 100, 50; 2: angle 25°, lengths 10, 50; angle 5° lengths 100,50) (18). For each candidate connectome, a probabilistic tracking algorithm (iFOD2) was used with a step size of 1 mm, a minimum length of 10 mm. The three candidate connectomes were then concatenated into a single ensemble connectome (ii) Spherical-deconvolution Informed Filtering of Tractograms (SIFT) to improve the quantitative accuracy of the ensemble connectome by filtering out streamlines that do not align with the FODs, ensuring that streamline densities match the FOD lobe integral. The resulting ensemble connectome retained 500,000 streamlines. (iii) AFQ (3) to segment and refine the left and right arcuate fasciculus from the resulting ensemble connectome of each neonate. Mean tract mean diffusivity (MD), fractional anisotropy (FA), and relaxation rate (R1) were calculated for the core of the tract, defined by the same ROIs used for tract segmentation of the left and right arcuate fasciculi.

Supplemental Tables

Supplemental Table S1. Trial intervention participant characteristics and intervention descriptives for the final sample with available diffusion MRI data.

| | **Treatment** | **Control** | **t or $X^2$** |
|---|---|---|---|
| **Participant characteristics** | *n* = 19 | *n* = 14 | |
| GA at birth (wk) | 29.7 (2.3) | 29.8 (2.1) | 0.22 |
| Birthweight (g) | 1364.2 (418.3) | 1389.4 (382.1) | 0.18 |
| SES[1] | 47.2 (17.5) | 45.3 (15.2) | -0.32 |
| Male (%) | 11 (57.79%) | 6 (42.9%) | 0.73 |
| Mechanical ventilation (days) | 1.4 (4.3) | 1.8 (4.3) | 0.24 |
| Antenatal corticosteroids (%) | 17 (89.5) | 13 (92.9) | 0.11 |
| Apgar at 1 min | 6.3 (2.4) | 5.4 (2.3) | -1.05 |
| Apgar at 5 min | 8.0 (1.4) | 8.0 (1.1) | 0. |
| **Intervention descriptives** | | | |
| PMA at start of intervention | 33.6 (0.8) | 34.0 (0.9) | 0.66 |
| Total days of intervention | 15.8 (8.7) | 16.2 (7.2) | 0.15 |
| PMA at MRI | 36.1 (1.3) | 36.0 (1.2) | 0.28 |
| Parent visitation during intervention (% days) | 70.4 (0.2) | 73.1 (0.2) | 0.41 |
| Voice recording in English (%) | 15 (78.9) | 12 (85.7) | 0.25 |

Supplemental Table S2. Pearson correlations between primary (MD) and secondary (FA, R1) outcomes of white matter microstructure.

| | Diffusion Scan 1 *Pearson's r* (95% CI) | | Diffusion Scan 2 *Pearson's r* (95% CI) | |
|---|---|---|---|---|
| | Left Arcuate | Right Arcuate | Left Arcuate | Right Arcuate |
| MD-FA | -0.52 (-0.75 - -0.18) | -0.75 (-0.88 - -0.53) | -0.87 (-0.94 - -0.69) | -0.68 (-0.86 - -0.36) |
| MD-R1 | -0.82 (-0.92- -0.63) | -0.85 (-0.93 – -0.67) | -0.78 (-0.91 - -0.51) | -0.74 (-0.89 - -0.43) |
| FA-R1 | 0.64 (0.32 - 0.83) | 0.79 (0.57 - 0.91) | 0.69 (0.35 - 0.87) | 0.56 (0.14 - 0.80) |

MD= mean diffusivity; FA = fractional anisotropy, CI = confidence interval

Supplemental Figure

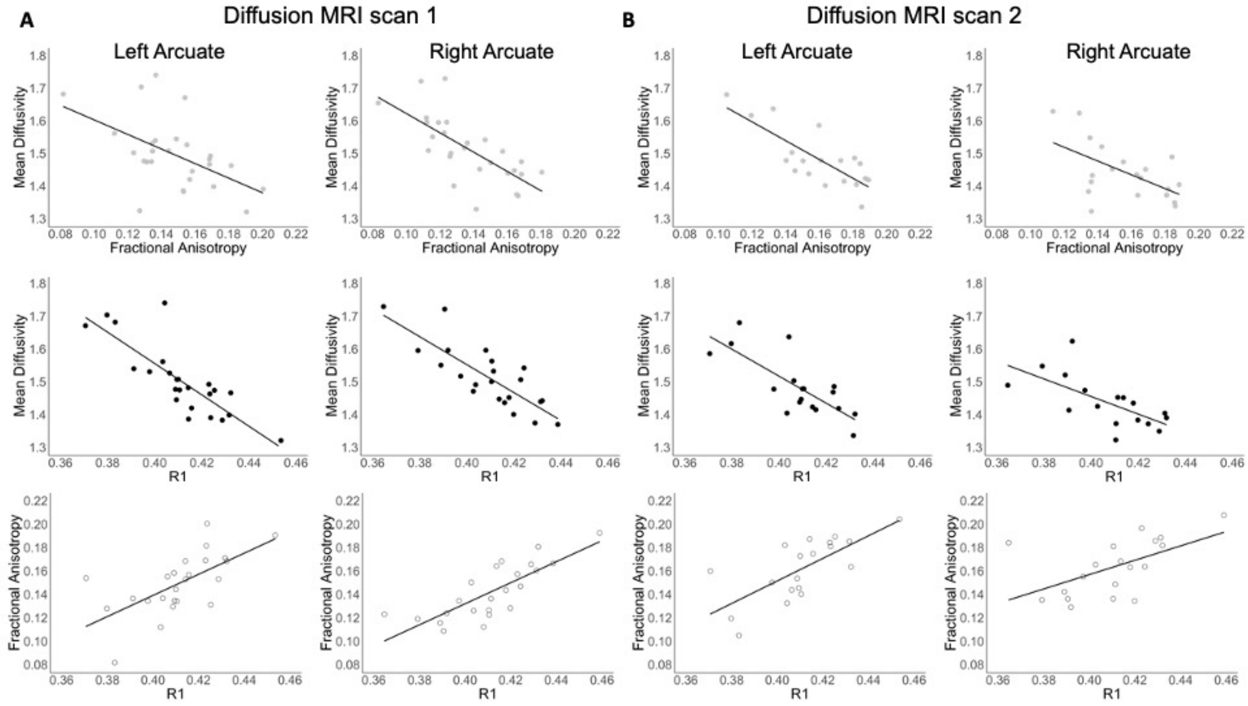


Supplemental Figure S1. Scatter plots depicting associations between primary (MD) and secondary outcome measures (FA, R1) of white matter microstructure. Panel A corresponds to diffusion MRI metrics MD and FA obtained from diffusion MRI scan 1 (bvalue = 700) and R1 obtained from the quantitative T1 relaxometry scan. Panel B corresponds to diffusion MRI metrics MD and FA obtained from diffusion MRI scan 2 (bvalue = 1500) and R1 obtained from the quantitative T1 relaxometry scan.
